# Evaluation of the Access Bio CareStart™ rapid SARS-CoV-2 antigen test in asymptomatic individuals tested at a community mass-testing program in Western Massachusetts

**DOI:** 10.1101/2021.06.17.21259109

**Authors:** Sara Suliman, Wilfredo R. Matias, Isabel R Fulcher, Francisco J. Molano, Shannon Collins, Elizabeth Uceta, Jack Zhu, Ryan M. Paxton, Sean F. Gonsalves, Maegan V. Harden, Marissa Fisher, Jim Meldrim, Stacey Gabriel, Molly F. Franke, Deborah T. Hung, Sandra C. Smole, Lawrence C. Madoff, Louise C. Ivers

## Abstract

**Background:** Point-of-care antigen-detecting rapid diagnostic tests (RDTs) for Severe Acute Respiratory Syndrome Coronavirus 2 (SARS-CoV-2) represent a scalable tool for SARS-CoV-2 infections surveillance. Data on their performance in real-world community settings is paramount for their implementation.

**Method:** We evaluated the accuracy of CareStart™ COVID-19 Antigen test (CareStart) in a testing site in Holyoke, Massachusetts. We compared CareStart to a SARS-CoV-2 reverse transcriptase quantitative polymerase chain reaction (RT-qPCR) reference, using anterior nasal swab samples. We calculated the sensitivity, specificity, and expected positive and negative predictive values at different SARS-CoV-2 prevalence estimates.

**Results:** We performed 666 tests on 591 unique individuals. 573 (86%) were asymptomatic. There were 52 positive tests by RT-qPCR. The sensitivity of CareStart was 49.0% (95% Confidence Interval (CI): 34.8 – 63.4) and specificity was 99.5% (95% CI: 98.5 – 99.9). Among positive RT-qPCR tests, the median cycle threshold (Ct) was significantly lower in samples that tested positive on CareStart. Using a Ct ≤ 30 as a benchmark for positivity increased the sensitivity to 64.9% (95% CI: 47.5 – 79.8).

**Conclusions:** CareStart has a high specificity and moderate sensitivity. The utility of RDTs, such as CareStart, in mass implementation should prioritize use cases in which a higher specificity is more important.

## Introduction

The Coronavirus disease 2019 (COVID-19) pandemic caused by the Severe Acute Respiratory Syndrome Coronavirus 2 (SARS-CoV-2) is the most significant infectious disease pandemic in the last century (1, 2). In addition to preventive measures such as social distancing, mask wearing, and vaccination, pillars of pandemic control rely on tools to rapidly identify cases and monitor transmission (3). Molecular testing methods based on reverse transcription quantitative polymerase chain reactions (RT-qPCR) remain the backbone of many testing programs globally (4). However, RT-qPCR-based testing is heavily influenced by supply chain restrictions, need for trained personnel and central laboratories, and relatively long turnaround times, particularly in resource-constrained settings (5). Therefore, it is still challenging to scale up RT-qPCR tests for population surveillance and the timely detection of the large proportion of asymptomatic SARS-CoV-2-infected carriers (6). Rapid detection of SARS-CoV-2 infected individuals allows for faster clinical intervention and implementation of public health measures such as isolation and contact-tracing, to prevent forward transmission (7).

Rapid antigen-detecting diagnostic tests (RDTs) for COVID-19, many of which can yield actionable results in turnaround times often below 20 minutes, require little laboratory capacity, and can be performed easily by non-laboratory personnel (8). Furthermore, decentralized access to RT-qPCR testing remains sparse in resource-constrained communities (9). The low cost of antigen-detecting RDTs, short turnaround times and ease of use make them excellent candidates to increase their accessibility for large-scale implementation in varied community settings (10).

Since the pandemic’s onset, several antigen-detecting RDTs have been developed for the detection of SARS-CoV-2 (11). Many of the antigen-detecting RDTs received Emergency Use Authorization (EUA) approvals by the Food and Drug Administration (FDA) (12). However, evaluations to receive EUA were performed by demonstrating accuracy in symptomatic individuals only (13). This narrow indication for antigen-detecting RDTs raises their limited utility in detecting SARS-CoV-2 infection in asymptomatic carriers. In fact, several independent evaluations demonstrate the decreased sensitivity of antigen-detecting RDTs in asymptomatic RT-qPCR positive individuals compared to those with symptoms (14–17). In the United States, studies thus far have focused on 3 RDTs: Quidel Sofia™ (8, 18), BD Veritor™ (13, 19) and Abbott BinaxNOW™ (15, 16, 20). On March 31^st^, 2021, the FDA also authorized these tests for home use, raising concerns about misinterpretation of false negative results (21). Therefore, evidence to establish their performance characteristics to guide their implementation in real-world settings is even more urgent now.

In this study, we evaluated the Access Bio CareStart™ COVID-19 RDT (CareStart), a chromatographic antigen-detecting lateral flow immunoassay that received EUA by the FDA (12, 17). We evaluated CareStart in asymptomatic and mildly symptomatic individuals presenting for routine testing at one of the ‘Stop the Spread’ free community testing sites in Holyoke, Massachusetts. Public health messaging for testing at these community testing sites targeted asymptomatic individuals. We evaluate the sensitivity, specificity, and positive (PPV) and negative predictive values (NPV) as a function of different prevalence scenarios.

## Methods

### Study Population and Ethical Approval

This was a prospective evaluation using convenience sampling of asymptomatic and mildly symptomatic individuals presenting for routine testing for COVID-19. The study was performed between January 6 and February 26, 2021, at the Holyoke “Stop the Spread” walk-up testing site, a free Massachusetts public testing program, which targets asymptomatic individuals (https://www.transformativehc.com/stopthespread.html). The testing site was open three days a week. Individuals who presented to the site during testing hours were approached by our research staff who explained the nature of the study, risks, benefits, and answered any questions before inviting individuals to participate in the study. Verbal consent was obtained from participants to collect a second anterior nasal swab as well as from guardians of minors below 18 years of age, from whom verbal assent was also obtained. The participants were treated in accordance with Good Clinical Practice guidelines and the Declaration of Helsinki. The study protocol was approved by the Partners Institutional Review Board (Protocol ID: 2020P003892).

### Study intake and data collection

After enrollment in the study, our study staff implemented an intake questionnaire capturing information on participant demographics, presence or absence of symptoms based on case definitions from the Council for State and Territorial Epidemiologists (22): cough, sore throat, chills, shortness of breath, fever, muscle aches or soreness, nausea, vomiting or diarrhea, decreased sense of smell or taste, loss of appetite, general weakness or fatigue, or headaches. The survey also captured prior COVID-19 testing and potential exposures. Each test was assigned a unique anonymous ID. Data collected was inputted into a secure Research Electronic Data Capture (REDCap) database on encrypted tablets. We used the demographic information and specimen numbers to match the RDTs result with the RT-qPCR data collected at the Broad Institute Clinical Research Sequencing Platform (CRSP) as performed in other studies (15, 23).

### Swab collection procedure

The sample was collected by trained personnel at the city testing site. We used dry anterior nasal (AN) swabs: Puritan 6” Sterile Standard Foam Swab with Polystyrene Handle (Puritan, Guilford). Both anterior nares were swabbed 2 times (5 rotations in each nostril), once for RT-qPCR testing and once for the RDT sample. For practical reasons, the swabs for RT-qPCR and RDT were not always collected in the same order. Both samples were placed inside closed test tubes. The RT-qPCR sample was transported to the Broad Institute at the Massachusetts Institute of Technology. The second anterior nasal swab sample was transported to a nearby testing station and the RDT was performed within an hour of sample collection. The RT-qPCR testing results were interpreted according to the publicly available rubric for the Broad Institute COVID-19 testing program: https://sites.broadinstitute.org/safe-for-school/result-code-information.

### Rapid test procedure

The CareStart device came with instructions for use and diagrams. The study staff received a one-hour training prior to the study and practiced the RDT on positive and negative control samples provided in the kit. One operator performed the test at a workstation following the CareStart manufacturer’s instructions for use (IFU) (24), took pictures of the tests, read the result as positive or negative, and captured into the electronic data entry forms. Participants with a positive RDT were contacted by phone per request from the Department of Public Health within a twenty-four-hour period, informed of their result, and advised to isolate until they received their RT-qPCR result.

### Reference RT-qPCR standard

The gold standard reference used was the SARS-CoV-2 RT-qPCR laboratory developed test through the Broad Institute CRSP, which is approved by the FDA under EUA. The test provides two cycle threshold (Ct) values, one for the nucleocapsid (N2) gene, and one for an internal positive control RNaseP gene. We compared the sensitivity of CareStart against both the qualitative binary RT-qPCR results and the Ct values of the N2 gene amplification reaction, as previously described (17).

### Statistical analyses

We calculated sensitivity, specificity, PPV and NPV of the RDT from 2 ⨯ 2 contingency tables using RT-qPCR as the gold standard reference. Tests with undetermined CareStart or RT-qPCR results were excluded from these calculations (n=35; 5.2% of all tests). Sensitivity and specificity were further stratified and compared by presence of symptoms and quantitative Ct values. Median Ct values were compared using the non-parametric unpaired Mann-Whitney *U* test. 95% Pearson-Clopper confidence intervals (CI) were calculated for sensitivity and specificity estimates. Since RDTs have been reported to have high accuracy among symptomatic individuals (8, 15–17), we also tested whether presence of symptoms would increase the sensitivity of the CareStart RDT. Statistical analyses were conducted using R V3.6.0 (R Core Team 2020).

## Results

We performed 666 CareStart RDTs from participants who provided verbal consent at the walk-up testing site **(Table 1 and Figure 1)**. The 666 tests performed were comprised of 591 unique participants with 60 participants by chance receiving more than one test, with a total of 75 tests performed in addition to the first test per participant **(Supplementary Table 1)**. Among the 591 participants, 47.9% were residents from Holyoke, as identified by their residential zip codes. Just over half the participants (51.9%) identified as female. The mean age was 38.1, and 44.7% of participants identified as Hispanic or LatinX **(Table 1A)**. Among Holyoke participants, 58.7% identified as Hispanic or LatinX **(Supplementary Table 2)**, in line with the reported demographics by the American Community Survey for 2019, which shows that 54% of Holyoke residents identified as LatinX. Participants who tested positive for the CareStart RDT were more likely to report at least one symptom than asymptomatic counterparts (41.9% vs. 12.2%; Chi-square p < 0.0001) **(Table 1B)**.

**Table 1A:** Demographics of unique study participants who enrolled in the CareStart Rapid Antigen Test evaluation at the Stop the Spread COVID-19 testing site in Holyoke, Massachusetts.

| **Demographics** | **Always Undetermined (N=3)** | **Always Negative (N=558)** | **Ever Positive (N=30)** | **Overall (N=591)** |
| --- | --- | --- | --- | --- |
| **Age (Years)**<br>Mean (SD)<br>Median [Min, Max] | 25.3 (3.51)<br>25 [22, 29] | 38.2 (17.9)<br>36 [1, 85] | 37.4 (15.4)<br>34.5 [16, 75] | 38.1 (17.8)<br>36 [1, 85] |
| **Sex**<br>Female<br>Male<br>Non-binary<br>Prefer not to answer<br>Not listed | 2 (66.7%)<br>1 (33.3%)<br>0 (0%)<br>0 (0%)<br>0 (0%) | 294 (52.7%)<br>236 (42.3%)<br>4 (0.7%)<br>24 (4.3%)<br>0 (0%) | 11 (36.7%)<br>14 (46.7%)<br>0 (0%)<br>4 (13.3%)<br>1 (3.3%) | 307 (51.9%)<br>251 (42.5%)<br>4 (0.7%)<br>28 (4.7%)<br>1 (0.2%) |
| **Lives in Holyoke zip code**<br>Yes<br>No | 3 (100%)<br>0 (0%) | 292 (52.3%)<br>266 (47.7%) | 13 (43.3%)<br>17 (56.7%) | 308 (52.1%)<br>283 (47.9%) |
| **Race/Ethnicity category**<br>Hispanic or LatinX<br>White Non-Hispanic<br>Asian Non-Hispanic<br>Black or African American Non-Hispanic<br>Native Hawaiian or Other Pacific Islander Non-Hispanic<br>Other Non-Hispanic<br>Two or more races Non-Hispanic<br>Prefer not to answer | 1 (33.3%)<br>2 (66.7%)<br>0 (0%)<br>0 (0%)<br>0 (0%)<br>0 (0%)<br>0 (0%)<br>0 (0%) | 248 (44.4%)<br>262 (47%)<br>5 (0.9%)<br>15 (2.7%)<br>1 (0.2%)<br>8 (1.4%)<br>4 (0.7%)<br>15 (2.7%) | 15 (50.0%)<br>14 (46.7%)<br>0 (0%)<br>0 (0%)<br>0 (0%)<br>0 (0%)<br>0 (0%)<br>1 (3.3%) | 264 (44.7%)<br>278 (47.0%)<br>5 (0.8%)<br>15 (2.5%)<br>1 (0.2%)<br>8 (1.4%)<br>4 (0.7%)<br>16 (2.7%) |
| **Repeat tester**<br>No<br>Yes | 3 (100%)<br>0 (0%) | 506 (90.7%)<br>52 (9.3%) | 22 (73.3%)<br>8 (26.7%) | 531 (89.8%)<br>60 (10.2%) |

**Table 1B:** Tester symptoms, exposure history and prior COVID-19 testing per each CareStart testing occurrence, including repeated tests from the same participants.

|  | **Negative**<br>**(N=631)** | **Positive**<br>**(N=31)** | **Undetermined**<br>**(N=4)** | **Overall**<br>**(N=666)** |
| --- | --- | --- | --- | --- |
| **Any symptoms** |  |  |  |  |
| No | 554 (87.8%) | 18 (58.1%) | 1 (25.0%) | 573 (86%) |
| Yes | 77 (12.2%) | 13 (41.9%) | 3 (75.0%) | 93 (14%) |
| **Exposure** |  |  |  |  |
| Confirmed or suspected | 196 (31.1%) | 17 (54.8%) | 4 (100%) | 217 (32.6%) |
| No exposure | 433 (68.6%) | 14 (45.2%) | 0 (0%) | 447 (67.1%) |
| Did not answer | 2 (0.3%) | 0 (0%) | 0 (0%) | 2 (0.3%) |
| **Prior COVID-19 test** |  |  |  |  |
| No | 110 (17.4%) | 9 (29.0%) | 2 (50.0%) | 121 (18.2%) |
| Yes | 519 (82.3%) | 22 (71.0%) | 2 (50.0%) | 543 (81.5%) |
| Missing | 2 (0.3%) | 0 (0%) | 0 (0%) | 2 (0.3%) |

**Figure 1:**
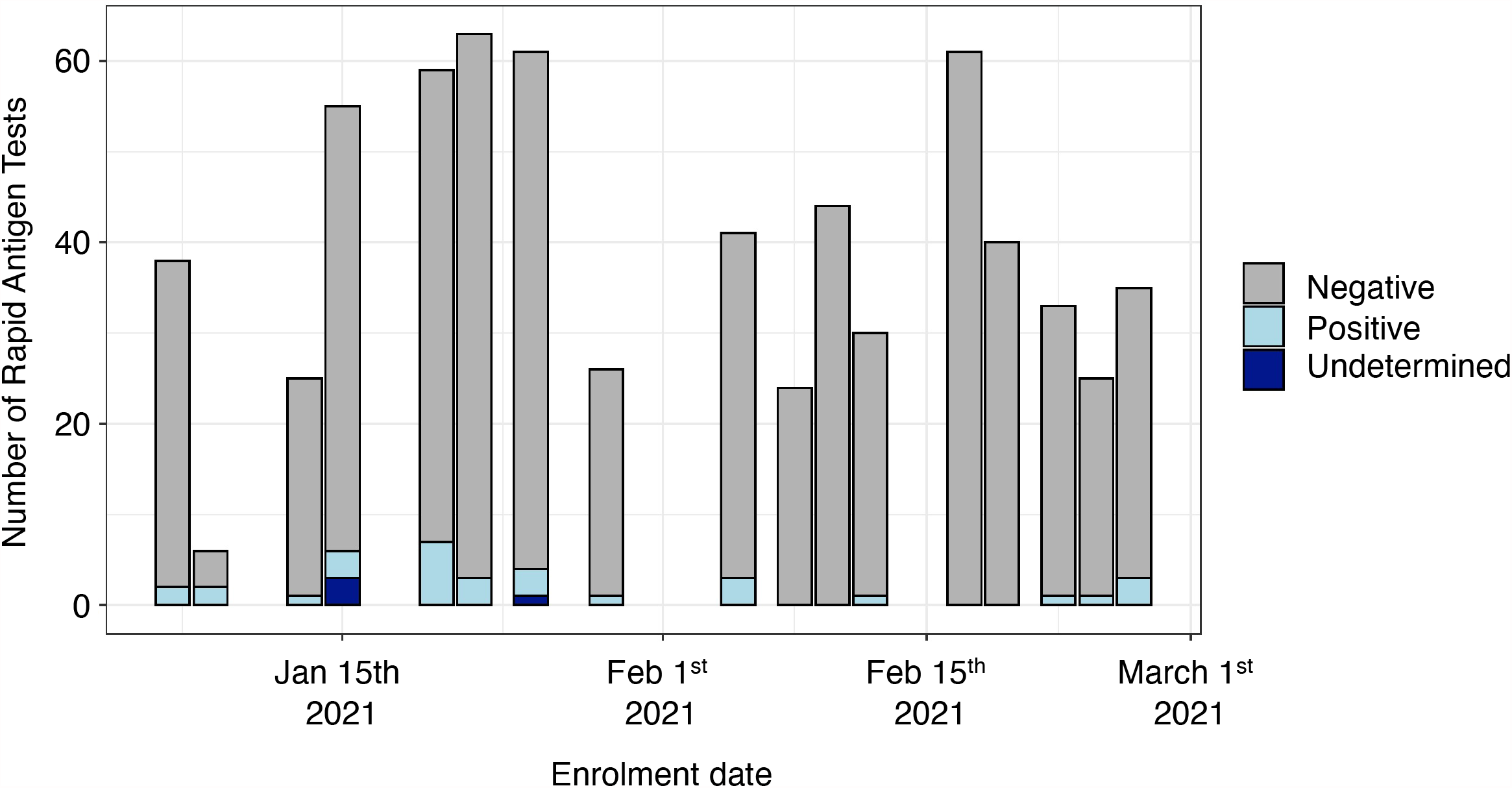
Number of CareStart rapid antigen test administered by date (n=666). The bar colors reflect the results of the rapid tests on different days.

The study staff evaluated the usability of the CareStart devices. All tests showed a positive control band, indicating they were valid. However, plastic in some vial caps was slightly malformed, making it difficult or impossible to properly cap the vials leading to 4 undetermined CareStart RDT results out of 666, since the operator could not load the RDT. Of the valid tests, we noted variable band intensities **(Figure 2)**. The positive test line was sometimes so faint that a flashlight was necessary to see it.

**Figure 2:**
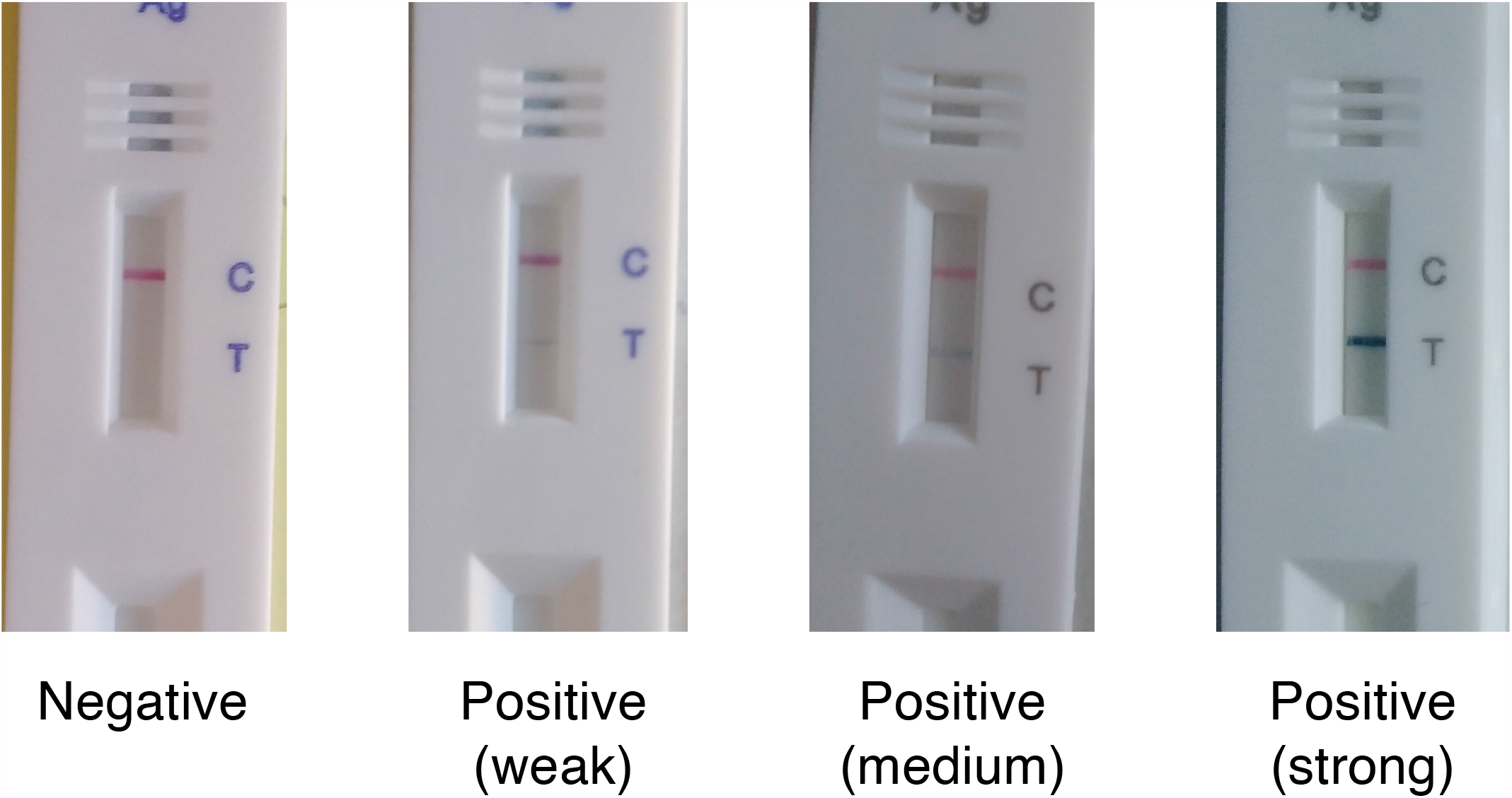
Examples of images of CareStart rapid test showing variable band intensities.

To determine the accuracy of the CareStart RDT, we calculated the concordance between the RDT and RT-qPCR **(Table 2)**. Using all RT-qPCR values below 40 as a positive reference, the sensitivity of the CareStart RDT was 48.1% (95% CI: 34.0%-62.4%), while the specificity was 99.0% (95% CI: 97.8%-99.6%) **(Table 3)**. Of the 666 visits, participants reported presence of symptoms 93 (14.0%) times **(Table 1B)**. Cough was the most reported (n=40, 6.0%) symptom, while loss of smell or taste, a more specific COVID-19 symptom, was only reported in 19 visits (2.9%) **(Supplementary Table 3)**. Due to the limited sample size, we only stratified individuals tested by presence (n=93) or absence (n=573) of symptoms to test the CareStart RDT accuracy as a function of symptoms. The sensitivity of CareStart RDT in symptomatic individuals was 46.4% (95% CI: 27.5%-66.1%), and the specificity was 100% (95% CI: 95%-100%) **(Supplementary Table 4A and 4B)**. In asymptomatic individuals, the sensitivity of the CareStart RDT was 52.2% (95% CI: 30.6%-73.2%), and the specificity was 99.4% (95% CI: 98.3%-99.9%) **(Supplementary Table 4C and 4D)**.

**Table 2:**
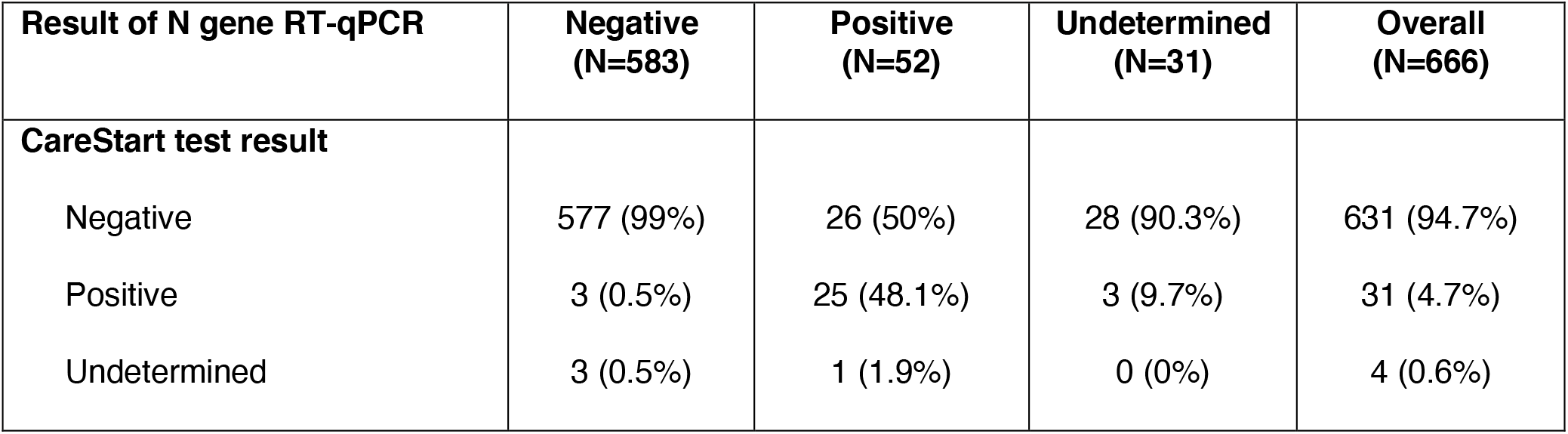
Concordance between CareStart Test Results and RT-qPCR test results.

| Result of N gene RT-qPCR | Negative<br>(N=583) | Positive<br>(N=52) | Undetermined<br>(N=31) | Overall<br>(N=666) |
| --- | --- | --- | --- | --- |
| **CareStart test result** |  |  |  |  |
| Negative | 577 (99%) | 26 (50%) | 28 (90.3%) | 631 (94.7%) |
| Positive | 3 (0.5%) | 25 (48.1%) | 3 (9.7%) | 31 (4.7%) |
| Undetermined | 3 (0.5%) | 1 (1.9%) | 0 (0%) | 4 (0.6%) |

**Table 3:** Performance characteristics of CareStart test results benchmarked against the RT-qPCR gold standard (excluding undetermined results).

|  | n | Total tests | Performance Characteristic | Estimate (%) | 95% Confidence Interval |
| --- | --- | --- | --- | --- | --- |
| **Rapid Test Results** |  |  |  |  |  |
| Positive | 25 | 51 | Sensitivity | 49.0% | (34.8% - 63.4%) |
| Negative | 577 | 580 | Specificity | 99.5% | (98.5% - 99.9%) |

Next, we used Ct values for amplification of the N2 target as a proxy for viral load, where higher Ct values reflected low viral loads, as previously reported (25). The Ct values of samples recorded as negative using the CareStart RDT were significantly higher than positive counterparts (Mann Whitney *U* p-value < 0.0001, **Figure 3)**. Therefore, we also performed a subset analysis where we only considered samples with a Ct < 30 as positive **(Supplementary Table 5A)**. Using this cut-off, the CareStart RDT sensitivity and specificity were 64.9% (95% CI: 47.5%-79.8%) and 99.3% (95% CI: 98.3%-99.8%), respectively **(Supplementary Table 5B)**. Although the CareStart RDT EUA does not indicate a specific Ct threshold for the positivity of the comparator RT-qPCR (24), these data suggest that applying a more stringent Ct value threshold moderately improves the sensitivity of the CareStart RDT.

**Figure 3:**
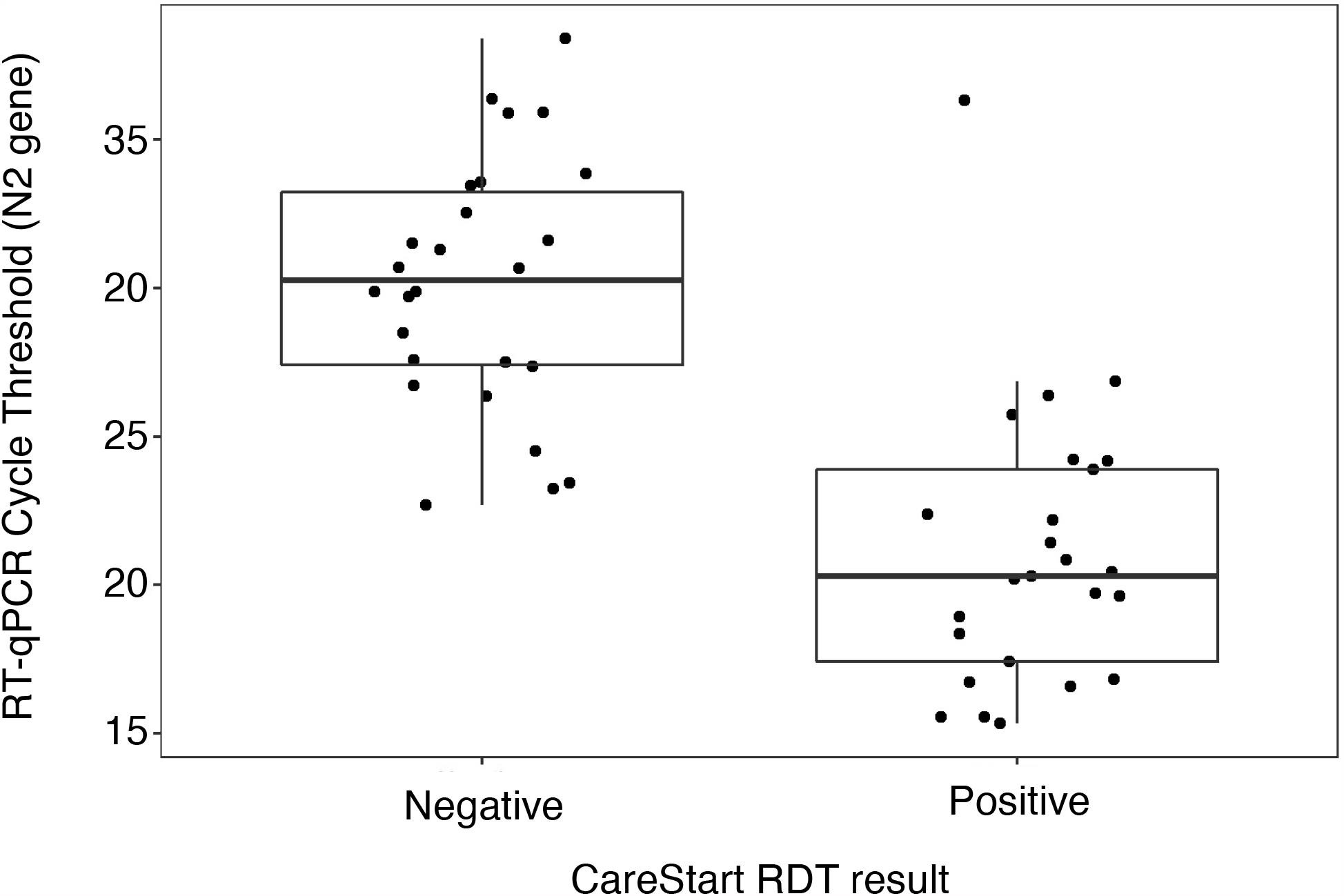
N2 gene RT-qPCR Cycle threshold (Ct) values corresponding to positive and negative CareStart rapid antigen test results for all RT-qPCR positive samples (n=52).

Positive and negative predictive values of diagnostic tests depend on the prevalence of infections in a population, where a higher prevalence increases the PPV at the expense of the NPV (26). We calculated the PPV and NPV values as a function of prevalence rates up to 10%, where the PPV steeply dropped in prevalence rates lower than 5% **(Figure 4)**. At a sensitivity of 49% and specificity of 99.5% **(Table 3)**, the PPV of CareStart was 49.7% at a SARS-CoV-2 infection prevalence of 1%, and 91.6% at a prevalence of 10%. In contrast, the NPV was 99.5% at a prevalence of 1%, and 94.6% at a prevalence of 10%.

**Figure 4:**
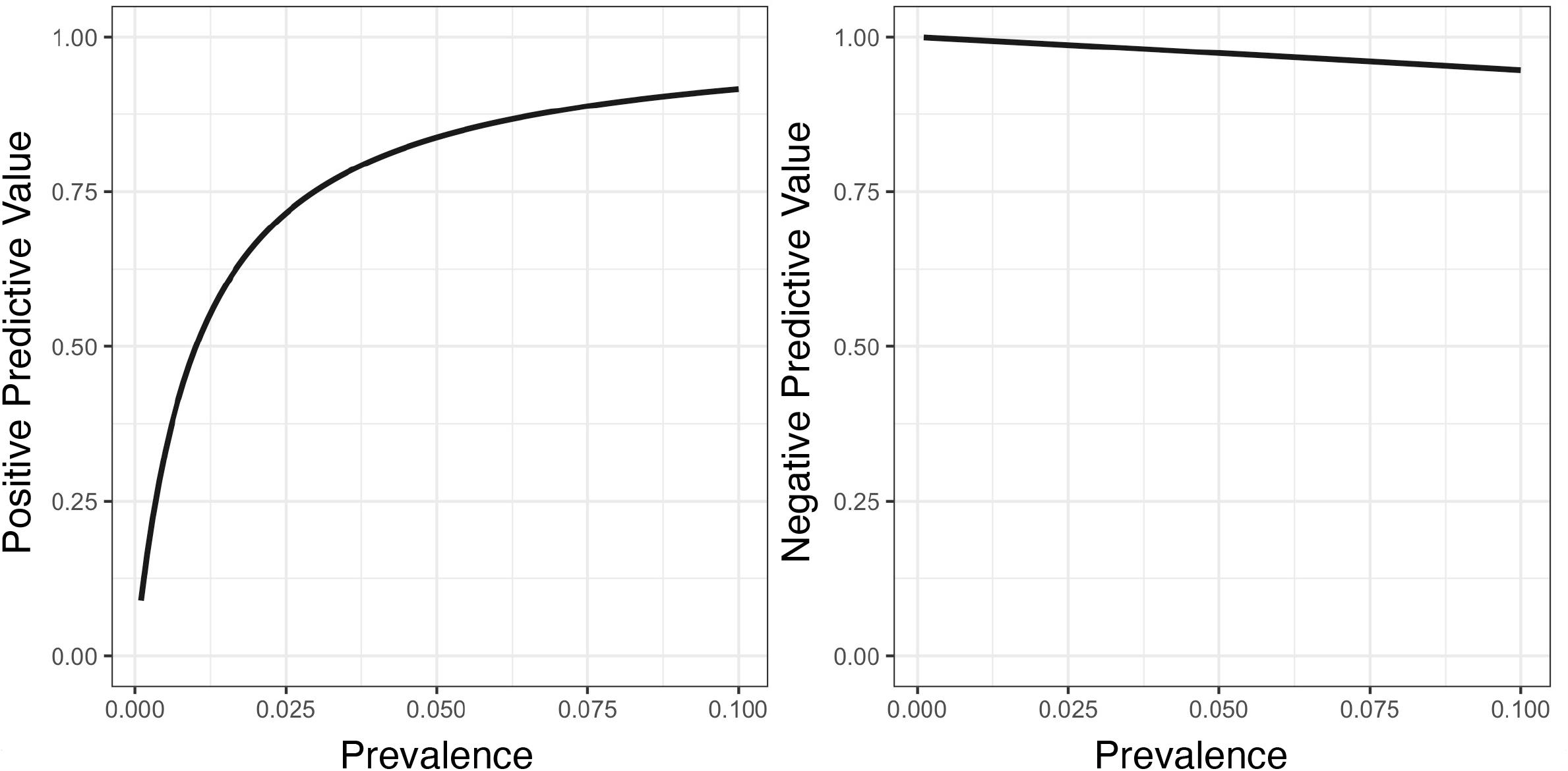
Calculated positive (left) and negative predictive values (right) based on the CareStart performance characteristics and different prevalence estimates of SARS-CoV-2 infections.

Finally, our cohort included individuals who presented to the testing site multiple times, who had at least one positive RT-qPCR test result. Therefore, we performed an exploratory analysis of their longitudinal test results **(Supplementary Table 1 and Figure 5)**. We enrolled 5 participants who converted from a negative to positive on RT-qPCR tests, all of which were accurately detected as positive by the RDT. Two participants with both positive RT-qPCR and RDT test results reverted to negative test results on both platforms. However, one participant converted from a positive to negative RDT test result but was detected as positive by the RT-qPCR on the second test, which was conducted in less than a week.

**Figure 5:**
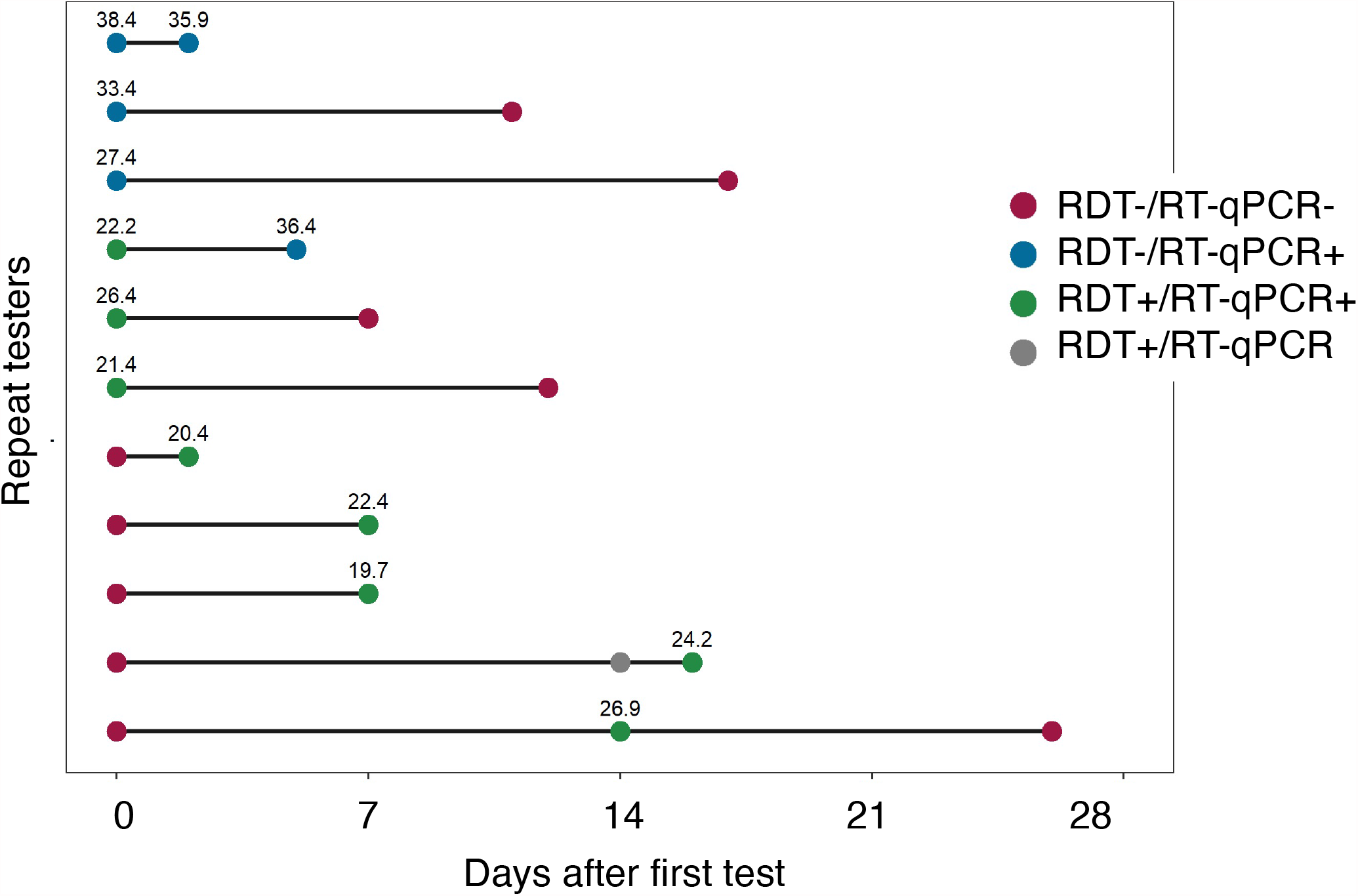
Individuals who enrolled in the study multiple times and had at least one positive gold standard RT-qPCR reference (n=11). The point colors reflect the different combinations of RT-qPCR and CareStart rapid test results. The numbers above the point correspond to Ct values of the RT-qPCR.

## Discussion

Antigen-detecting RDTs could provide a scalable and affordable alternative to molecular tests (27). In this study, we present a prospective evaluation of the CareStart antigen-detecting RDT for SARS-CoV-2 detection in a walk-up community COVID-19 testing site in Holyoke, Western Massachusetts. The SARS-CoV-2 testing gap refers to the disparity between where communities rank with respect to numbers of tests performed per 100,000 individuals, and where they rank with respect to SARS-CoV-2 positivity rates in each state (28). Equitable implementation of testing should lead to higher numbers of tests performed per capita in communities with higher proportions of COVID-19 cases (29). In the state of Massachusetts, the SARS-CoV-2 testing gap, was higher in communities with a high social vulnerability index, including Holyoke, highlighting the socioeconomic disparities underlying access to COVID-19 testing (28). Thus, scaling up affordable tests such as antigen-detecting RDTs can help fill this testing gap, following evaluation of their accuracy and implementation feasibility.

Compared to a RT-qPCR-based test, we found a much lower sensitivity (49.0%) than what was reported in the FDA package insert (87.2%), which was restricted to 39 symptomatic individuals within 5 days of symptom onset (24). However, this sensitivity was consistent with the reported sensitivity of CareStart in asymptomatic individuals recruited in a recent study at Lawrence General Hospital in Massachusetts (84.8%) (17). Compared to the Abbott BinaxNOW RDT, which has been validated in several studies including a recent study in Massachusetts, the sensitivity of the CareStart RDT was lower overall and when using a Ct positivity cutoff of ≤ 30 (15). The specificity of both tests was comparable at nearly > 99 %. Consistent with several studies, the sensitivity of antigen detecting RDTs was modest in individuals with no or mild symptoms (8, 14-16, 20). Although viral loads (30) and recovery of viable virus (31) peak in the early days following symptom onset, transmission by asymptomatic individuals remains a main driver of the pandemic (6), especially since absence of symptoms removes the prompt to quarantine. Therefore, implementation of antigen-detecting RDTs needs to weigh the benefits of rapid detection of SARS-CoV-2-infected individuals with the lower sensitivity of these tests in asymptomatic carriers (27). On the other hand, the high specificity of these tests reduces the probability of false positive SARS-CoV-2 test results, that would have otherwise led to restrictions and inconveniences that may interfere with the livelihood of these individuals.

Evaluation of RDTs against a highly sensitive gold standard such as RT-qPCR are important to define performance characteristics. The RT-qPCR cycle threshold (Ct) values had a clear impact on the sensitivity of the CareStart RDT, where concordant results showed lower Ct values. In contrast, discordant results had higher Ct values, a proxy for lower viral load, as reported in other evaluations (15, 17, 32). Consistent with this, the CareStart RDT sensitivity improved with a RT-qPCR positivity Ct cut-off of < 30. These data suggest that the CareStart RDT positivity meant higher viral load, which correlates with infectivity of cells *in vitro* (33), and likely transmissibility. Individuals vaccinated with the Pfizer/BioNtech BNT162b2 vaccine, with breakthrough SARS-CoV-2 infections have been reported to show higher Ct values post-vaccination than unvaccinated counterparts, which correlates with lower viral loads (34). The implication, however, is that vaccinated individuals with breakthrough SARS-CoV-2 infections, will be more difficult to detect by antigen-detecting RDT with similar performance characteristics to CareStart. In a study in skilled nursing facility in Chicago, few and mostly asymptomatic break-through infections were identified post-vaccination (35). However, two were hospitalized and one died of COVID-19, suggesting that false negative diagnosis of SARS-CoV-2 infections are likely in postvaccination breakthrough infections (35). Therefore, it is crucial to evaluate the accuracy of RDTs in populations with high vaccination rates. Furthermore, false negative results of antigen-detecting RDTs may be caused by SARS-CoV-2 variants, which may be more transmissible (36). In laboratory studies, lower RT-qPCR Ct values were shown to correlate with increased recovery of viable virus (31, 37). However, epidemiological studies to support the higher contagiousness of individuals with lower Ct values are still lacking. Furthermore, our study shows overlapping RT-qPCR Ct values that could be either detected as positive or negative by the CareStart RDTs. Furthermore, 4 out of 26 negative RDT tests results corresponding to a positive RT-qPCR test result showed relatively low RT-qPCR Ct values (Ct < 25), indicating that false negative RDT results can occur even at high viral loads. Thus, to understand the public health implications of test results, it is imperative to definitively determine the infectiousness of individuals with a positive RT-qPCR, but negative RDT result, even if those negative calls only occurred occasionally.

Although our sample of repeat testers was limited, it suggested that individuals who recently converted from negative to positive RT-qPCR test, i.e. recently acquired SARS-CoV-2 infection, were easily detectable by the CareStart RDT. Recently infected individuals have been shown to be more contagious (25). Samples from these recently infected individuals had low Ct values, and were thus more likely to transmit the virus (33). Alternatively, the one participant with a “false negative” RDT result after testing positive twice with the RT-qPCR, showed a relatively high RT-qPCR Ct value of 36.4 on the second discordant result, suggesting lower contagiousness. Repeat testing may be influenced by school or work requirements and may reflect a population with a higher susceptibility to exposure such as health care workers. Therefore, this limited sample supports the potential usefulness of serial rapid antigen testing in detecting recent infections to implement containment measures and merits further study (38). Importantly, the public health benefits of serial testing with RDTs should be studied further.

There are several advantages to rapid testing. CareStart has a lower turnaround time than RT-qPCR, including 10 minutes to run the test on site where most of the test results were visible in 3-4 minutes. Infected individuals can be notified of their status and begin isolation immediately, resulting in less transmission to contacts compared to tests with longer turnaround times. In addition, using anterior nasal swabs instead of the internationally accepted gold standard nasopharyngeal swabs increases the acceptability of testing. However, it may also compromise the sensitivity of antigen detecting RDTs, although a recent evaluation of swab types disputes this possibility (39). The test is simple, and requires little training to be performed, making it more amenable to self-administration and home testing. The price of RDTs is gradually decreasing to become more affordable. Some individuals might test by RDT who would otherwise not be tested at all. Therefore, a formal assessment of the public health impacts of RDTs should be conducted in future studies.

A few factors may have compromised the performance characteristics of CareStart in this study. We had limited control over or ability to monitor the order by which the two bilateral nasal swabs were collected because of embedding the study in a ‘real world’ testing program. It is possible that performing the PCR swab first may decrease the available viral load for the antigen test. However, a recent evaluation of the Abbott BinaxNOW suggested that the order of swabs had little impact on the test result (20). The tests were analyzed indoors to minimize temperature fluctuations. However, the environmental temperature was not systematically monitored in this semi-indoor setting (foyer of the War Memorial Building, Holyoke, MA). Since the study was performed during the winter in Massachusetts, the tests were generally conducted at cold temperatures, which could impact the RDT performance (15, 40). These results are, hence, reflective of a real-world evaluation scenario, where lower temperatures and seasonal fluctuations are expected in regions such as Massachusetts. Finally, study was conducted at the time when vaccination against SARS-CoV-2 was low, which may affect the generalizability of these findings. In conclusion, RDTs such as CareStart, can support SARS-CoV-2 testing efforts in minimally or asymptomatic individuals. However, the impact of the limited sensitivity of these tests on their positive predictive values reinforce caution. The moderate sensitivity of these tests means that some potentially infectious individuals may be classified as SARS-CoV-2-negative. Therefore, implementing RDTs for travel, home testing, or to guide re-openings of schools and workplaces should be interpreted with caution and the utility of RDTs in each of these use cases should be carefully evaluated. Furthermore, implementation studies to analyze their usefulness and acceptability by both users and providers are necessary.

## Data Availability

Summary data, without any identifying individual information is reported in the manuscript's supplementary tables.

## Acknowledgements

The Holyoke Board of Health was instrumental in coordinating study logistics with Fallon Ambulance Services who performed the swabs for RT-qPCR. The MA Department of Public Health and Ginko Bioworks donated the Access Bio testing kits and anterior nares SteriPack swabs for this study. The study was supported by an award from the Massachusetts Life Sciences Center Accelerating Coronavirus Testing Solutions (A.C.T.S) and the Sullivan Family Foundation (LCI).

## Potential conflicts of interest

None.

---

**Supplementary Table 1:**
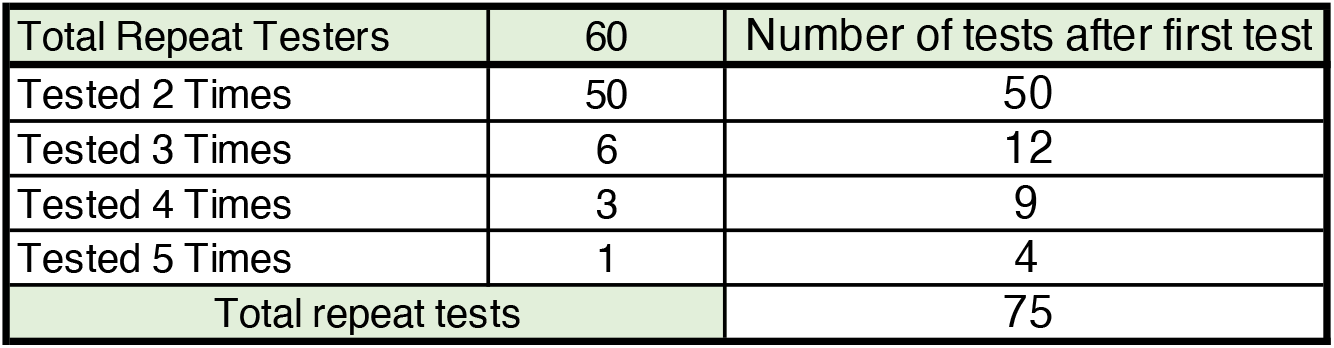
Study participants who enrolled multiple times for testing with both RT-qPCR and Access Bio CareStart.

| Total Repeat Testers | 60 | Number of tests after first test |
| --- | --- | --- |
| Tested 2 Times | 50 | 50 |
| Tested 3 Times | 6 | 12 |
| Tested 4 Times | 3 | 9 |
| Tested 5 Times | 1 | 4 |
| Total repeat tests |  | 75 |

**Supplementary Table 2:**
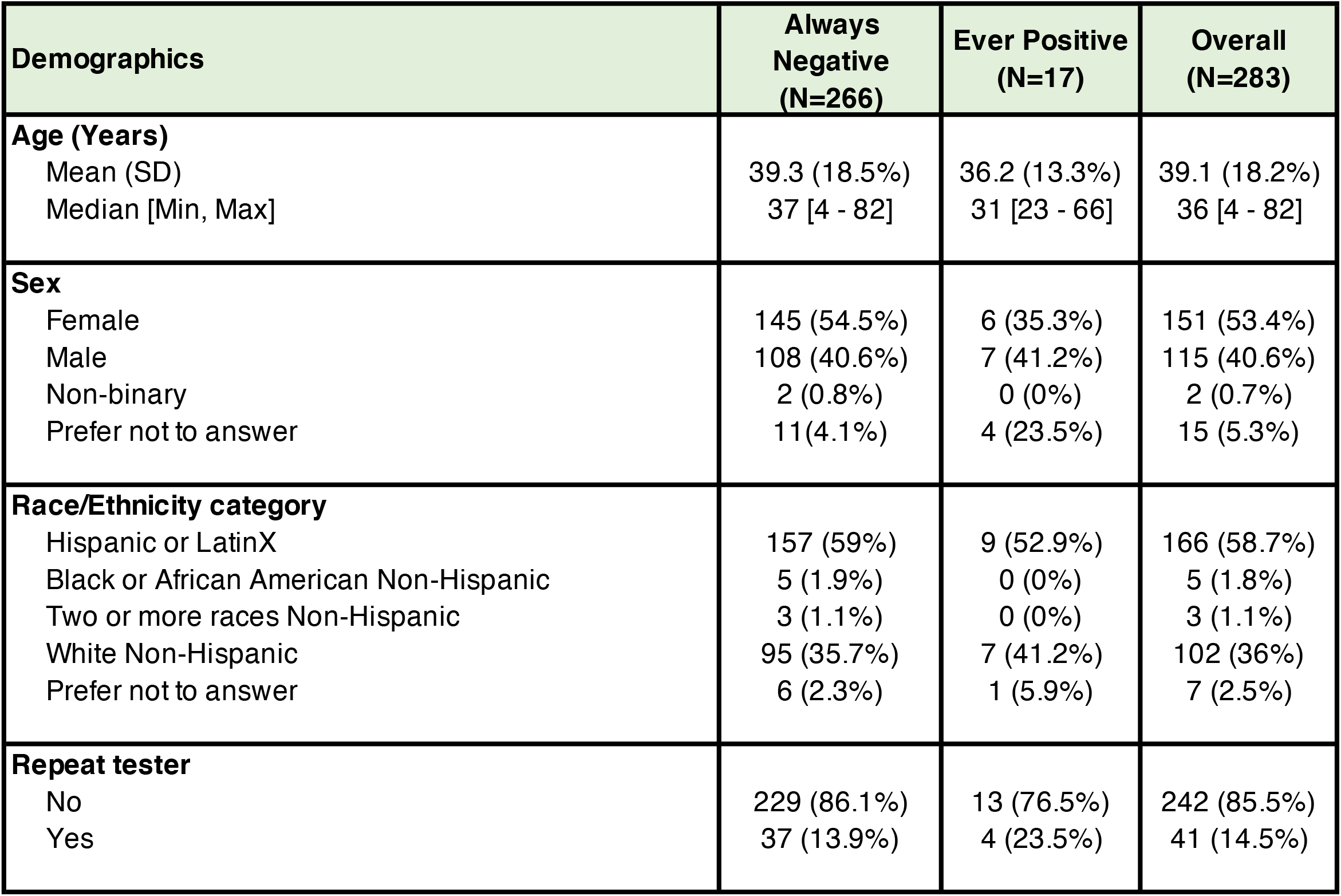
Demographics of study participants who are residents of Holyoke, Massachusetts (N=283)

| Demographics | Always Negative (N=266) | Ever Positive (N=17) | Overall (N=283) |
| --- | --- | --- | --- |
| **Age (Years)**<br>Mean (SD)<br>Median [Min, Max] | 39.3 (18.5%)<br>37 [4 - 82] | 36.2 (13.3%)<br>31 [23 - 66] | 39.1 (18.2%)<br>36 [4 - 82] |
| **Sex**<br>Female<br>Male<br>Non-binary<br>Prefer not to answer | 145 (54.5%)<br>108 (40.6%)<br>2 (0.8%)<br>11(4.1%) | 6 (35.3%)<br>7 (41.2%)<br>0 (0%)<br>4 (23.5%) | 151 (53.4%)<br>115 (40.6%)<br>2 (0.7%)<br>15 (5.3%) |
| **Race/Ethnicity category**<br>Hispanic or LatinX<br>Black or African American Non-Hispanic<br>Two or more races Non-Hispanic<br>White Non-Hispanic<br>Prefer not to answer | 157 (59%)<br>5 (1.9%)<br>3 (1.1%)<br>95 (35.7%)<br>6 (2.3%) | 9 (52.9%)<br>0 (0%)<br>0 (0%)<br>7 (41.2%)<br>1 (5.9%) | 166 (58.7%)<br>5 (1.8%)<br>3 (1.1%)<br>102 (36%)<br>7 (2.5%) |
| **Repeat tester**<br>No<br>Yes | 229 (86.1%)<br>37 (13.9%) | 13 (76.5%)<br>4 (23.5%) | 242 (85.5%)<br>41 (14.5%) |

**Supplementary Table 3:**
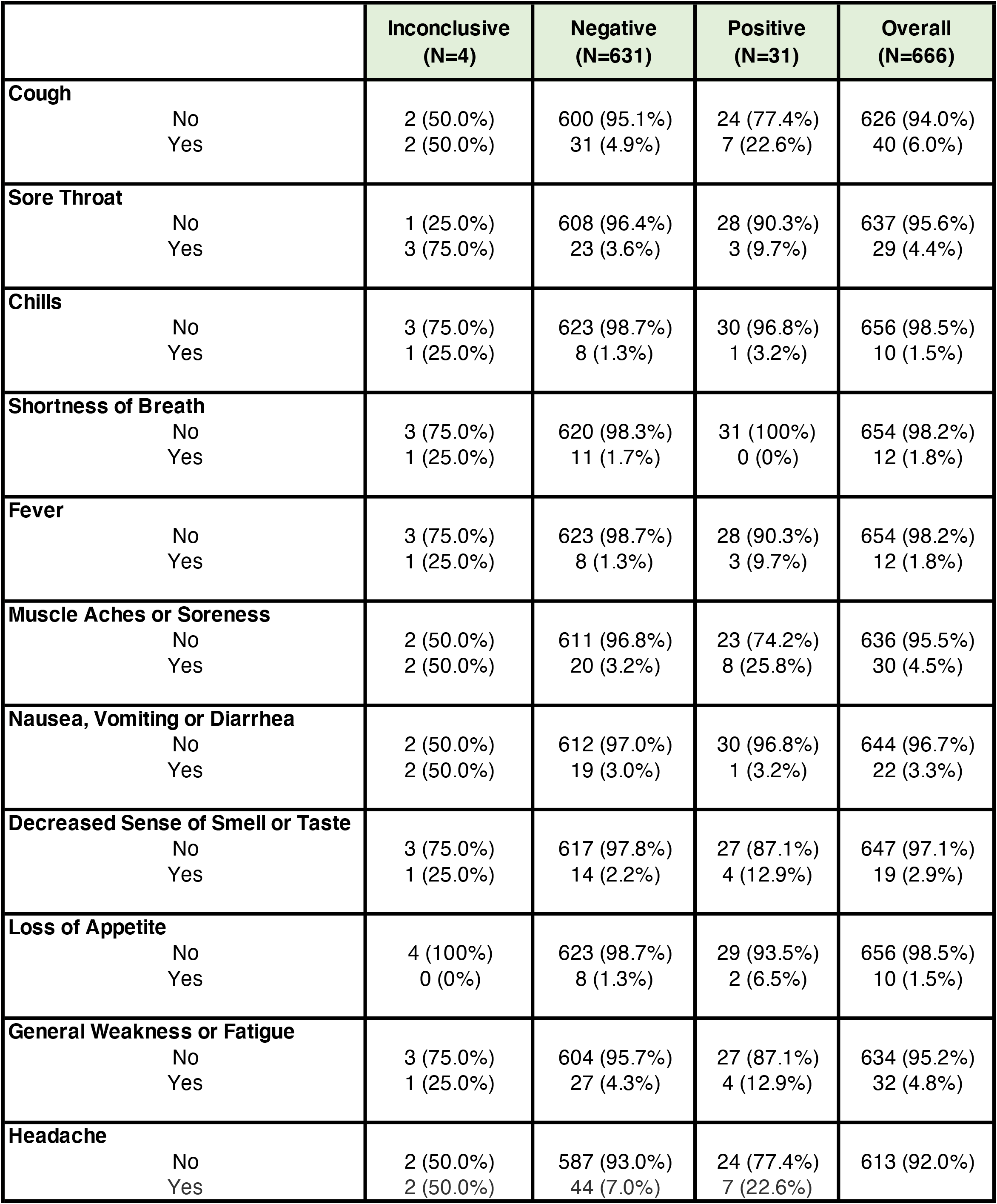
Symptoms reported at all visits.

|  | Inconclusive<br>(N=4) | Negative<br>(N=631) | Positive<br>(N=31) | Overall<br>(N=666) |
| --- | --- | --- | --- | --- |
| **Cough** |  |  |  |  |
| No | 2 (50.0%) | 600 (95.1%) | 24 (77.4%) | 626 (94.0%) |
| Yes | 2 (50.0%) | 31 (4.9%) | 7 (22.6%) | 40 (6.0%) |
| **Sore Throat** |  |  |  |  |
| No | 1 (25.0%) | 608 (96.4%) | 28 (90.3%) | 637 (95.6%) |
| Yes | 3 (75.0%) | 23 (3.6%) | 3 (9.7%) | 29 (4.4%) |
| **Chills** |  |  |  |  |
| No | 3 (75.0%) | 623 (98.7%) | 30 (96.8%) | 656 (98.5%) |
| Yes | 1 (25.0%) | 8 (1.3%) | 1 (3.2%) | 10 (1.5%) |
| **Shortness of Breath** |  |  |  |  |
| No | 3 (75.0%) | 620 (98.3%) | 31 (100%) | 654 (98.2%) |
| Yes | 1 (25.0%) | 11 (1.7%) | 0 (0%) | 12 (1.8%) |
| **Fever** |  |  |  |  |
| No | 3 (75.0%) | 623 (98.7%) | 28 (90.3%) | 654 (98.2%) |
| Yes | 1 (25.0%) | 8 (1.3%) | 3 (9.7%) | 12 (1.8%) |
| **Muscle Aches or Soreness** |  |  |  |  |
| No | 2 (50.0%) | 611 (96.8%) | 23 (74.2%) | 636 (95.5%) |
| Yes | 2 (50.0%) | 20 (3.2%) | 8 (25.8%) | 30 (4.5%) |
| **Nausea, Vomiting or Diarrhea** |  |  |  |  |
| No | 2 (50.0%) | 612 (97.0%) | 30 (96.8%) | 644 (96.7%) |
| Yes | 2 (50.0%) | 19 (3.0%) | 1 (3.2%) | 22 (3.3%) |
| **Decreased Sense of Smell or Taste** |  |  |  |  |
| No | 3 (75.0%) | 617 (97.8%) | 27 (87.1%) | 647 (97.1%) |
| Yes | 1 (25.0%) | 14 (2.2%) | 4 (12.9%) | 19 (2.9%) |
| **Loss of Appetite** |  |  |  |  |
| No | 4 (100%) | 623 (98.7%) | 29 (93.5%) | 656 (98.5%) |
| Yes | 0 (0%) | 8 (1.3%) | 2 (6.5%) | 10 (1.5%) |
| **General Weakness or Fatigue** |  |  |  |  |
| No | 3 (75.0%) | 604 (95.7%) | 27 (87.1%) | 634 (95.2%) |
| Yes | 1 (25.0%) | 27 (4.3%) | 4 (12.9%) | 32 (4.8%) |
| **Headache** |  |  |  |  |
| No | 2 (50.0%) | 587 (93.0%) | 24 (77.4%) | 613 (92.0%) |
| Yes | 2 (50.0%) | 44 (7.0%) | 7 (22.6%) |  |

**Supplementary Table 4A:** CareStart test results compared to RT-qPCR in symptomatic individuals only.

| CareStart Results | RT-qPCR Results |  |  |  |
| --- | --- | --- | --- | --- |
|  | Negative (N=61) | Positive (N=29) | Undetermined (N=3) | Overall (N=93) |
| Negative | 59 (96.7%) | 15 (51.7%) | 3 (100%) | 77 (82.8%) |
| Positive | 2 (3.3%) | 1 (3.4%) | 0 (0%) | 3 (3.2%) |
| Undetermined | 0 (0%) | 13 (44.8%) | 0 (0%) | 13 (14%) |

**Supplementary Table 4B:**
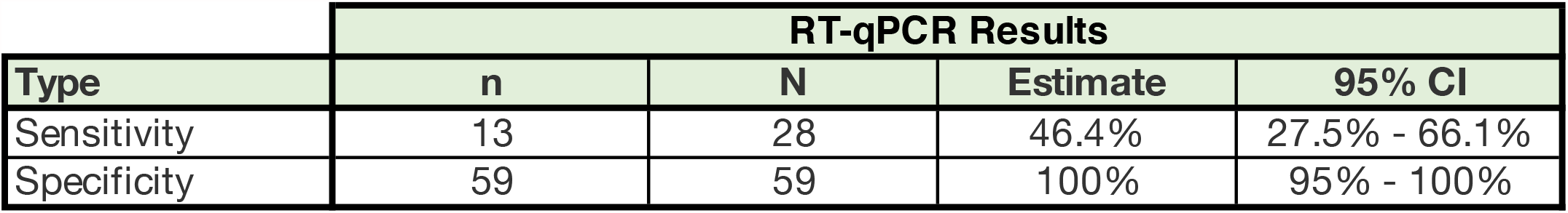
CareStart test result sensitivity and specificity in symptomatic individuals only.

**Supplementary Table 4C:** CareStart test results compared to RT-qPCR in asymptomatic individuals only.

| CareStart Results | RT-qPCR Results |  |  |  |
| --- | --- | --- | --- | --- |
|  | Negative (N=522) | Positive (N=23) | Undetermined (N=28) | Overall (N=573) |
| Negative | 518 (99.2%) | 11 (47.8%) | 25 (89.3%) | 554 (96.7%) |
| Positive | 3 (0.6%) | 12 (52.2%) | 3 (10.7%) | 18 (3.1%) |
| Undetermined | 1 (0.2%) | 0 (0%) | 0 (0%) | 1 (0.2%) |

**Supplementary Table 4D:** CareStart test result sensitivity and specificity in asymptomatic individuals only.

| Type | RT-qPCR Results |  |  |  |
| --- | --- | --- | --- | --- |
|  | n | N | Estimate | 95% CI |
| Sensitivity | 12 | 23 | 52.2% | 30.6% - 73.2% |
| Specificity | 518 | 521 | 99.4% | 98.3% - 99.9% |

**Supplementary Table 5A:** CareStart test results compared to RT-qPCR using Ct positivity threshold of < 30.

| CareStart Results | Negative (Ct >=30)<br>N=597 | Positive (Ct <30)<br>N=38 | Undetermined<br>N=31 | Overall N=666 |
| --- | --- | --- | --- | --- |
| Negative | 590 (98.8%) | 13 (34.2%) | 28 (90.3%) | 631 (94.7%) |
| Positive | 4 (0.7%) | 24 (63.2%) | 3 (9.7%) | 31 (4.7%) |
| Undetermined | 3 (0.5%) | 1 (2.6%) | 0 (0%) | 4 (0.6%) |

**Supplementary Table 5B:** CareStart test result sensitivity and specificity using Ct positivity threshold of < 30.

| Type | n | N | Estimate | 95% CI |
| --- | --- | --- | --- | --- |
| Sensitivity | 24 | 37 | 64.9% | 47.5%-79.8% |
| Specificity | 590 | 594 | 99.3% | 98.3% - 99.8% |

## Notes

### Competing Interest Statement

The authors have declared no competing interest.

### Author Declarations

Verbal consent was obtained from participants to collect a second anterior nasal swab as well as from guardians of minors below 18 years of age, from whom verbal assent was also obtained. The participants were treated in accordance with Good Clinical Practice guidelines and the Declaration of Helsinki. The study protocol was approved by the Partners Institutional Review Board (Protocol ID: 2020P003892).

